# Decoding imaginary handwriting trajectories with shape and time distortion loss for brain-to-text communication

**DOI:** 10.1101/2024.07.02.24309802

**Authors:** Guangxiang Xu, Zebin Wang, Kedi Xu, Junming Zhu, Jianmin Zhang, Yueming Wang, Yaoyao Hao

## Abstract

The potential to decode handwriting trajectories from brain signals has yet to be fully explored in clinical brain-computer interfaces (BCIs). One of challenges remains that the clinical BCIs mostly rely on imaginary movement due to motor deficit of the subject, which often leads to misalignment with neural activity and impedes accurate decoding. Here, we recorded intracortical neural signals from a paralyzed patient during imaginary handwriting of Chinese characters, from which the trajectories of handwriting were reconstructed and translated into texts using machine learning approach. We introduced an innovated decoding framework that incorporates a novel loss function, DILATE, to accommodate both shape and temporal distortions between movement and neural activity to account for the misalignment issue. Our method reconstructed closely resembled and human-recognizable handwriting trajectories, outperforming the conventional mean square error loss by 10% of recognition rate. Moreover, the new decoding framework enabled effective multi-day data fusion, resulting in further 15% enhancement. With a dynamic time warping approach, the recognition rate achieved up to 91.1% within a 1000-character database. Additionally, we applied our method to a previous subject who imaged handwriting of English letters, showcasing its capability for single-trail trajectory reconstruction and 13.5% higher recognition outcomes. Altogether, these findings demonstrated a new decoding scheme for BCIs that could accurately reconstruct the imaginary handwriting trajectory. This advancement paves the way for a universal brain-to-text communication system that is applicable to any written language, marking a significant leap forward in the field of neural decoding and BCI technology.

## Introduction

Over the past two decades, intracortical brain-computer interfaces (iBCIs) have emerged as a revolutionary tool that enables direct communication between the human brain and external devices [1-9]. The iBCIs have opened new avenues for individuals with motor deficits to interact more effectively with their surroundings, leading to extensive clinical trials being conducted worldwide [10].

Trajectory fitting is a well-established technique in BCIs, which translates movement intention into continuous motion of computer cursors or robotic devices [11, 12]. Most of the prior studies have employed mean squared error (MSE) loss to optimize the trajectory fitting algorithms, aiming to develop optimal decoders [13, 14]. However, MSE has its own inherent limitations when assessing predictions, particularly in scenarios with sharp variations [15-17]. While this limitation is not critical in simple trajectory tasks, such as center-out movements, it becomes pronounced in complex cases like cursive moving [18, 19] or handwritings [7, 20], where precise path following is required. Furthermore, in clinical BCI applications, there is often an absence of labeled movement to training the decoder, due to the motor impairment of the subject. In this case, an initial imaginary movement process was conducted following a guidance for the subject [7, 8]. These guidance movements are taken as the surrogate label to train an initial decoder, which is then further optimized during subsequent online control experiments. However, the subject’s thoughts may either precede or lag the guidance movements, leading to misalignment which results in less precise decoding models. The MSE method enforces a point-to-point alignment and distributes the loss across the entire trial to achieve a global minimum, which is not well-suited for situations with misalignment [21]. Therefore, there is a clear demand for a loss function capable of accounting for alignment with sharp variations to ensure accurate decoding in practical BCI settings.

The incorporation of handwriting paradigm into BCIs marks a substantial advancement in the field, enabling users to convert imagined handwriting movements into textual output. A seminal work in this domain was done by Willett et al. [7], which demonstrated the feasibility of classifying neural activity into English letters with an impressive 94.1% raw classification accuracy within a 30-character scope. This process was then efficiently transcribed into text at a rate of up to 90 characters per minute. These advancements highlighted the exceptional capabilities of a high-performance BCI [7]. However, the classification-based decoding scheme utilized in the study was specifically designed for Latin-based languages, which require discrimination of only a few dozen letters to form text. In contrast, non-Latin languages, such as Chinese, demand classification of thousands of distinct characters - a challenge that is currently beyond the capabilities of neural signal-based classification for BCIs. Therefore, there is an urgent need for an innovative decoding scheme that could reconstruct the trajectories of imagined handwritings, instead of just identities, to realize a universal brain-to-text system for any written languages. In this context, the overall shape of the handwriting trajectories, even if they include misalignment or inaccurate amplitude, is far more critical than merely the overall minimum MSE. This underscores the importance of developing a decoding approach that prioritizes the preservation of the trajectory’s overall shape, which is essential for accurate text translation across diverse languages.

In this study, we acquired intracortical neural signals from a patient during imaging of handwritings, with the goal to reconstruct the very trajectories of the imagined handwritings and recognize them as standard text. We introduced a novel distortion loss function (DILATE) [15], which incorporates both dynamic time warping (DTW) based shape matching along with a temporal term, to train the trajectory fitting model. Comparing with MSE, DILATE loss achieved high-fidelity of trajectory reconstruction and better recognition rate in both single-day and multi-day fusion decoding. Furthermore, the DILATE loss was also applied to the previous handwriting dataset for English letters [7] and demonstrated superior single-trial trajectory reconstruction and better recognitions. These advancements hold profound implications for the field of BCI, offering a promising new pathway towards more accurate handwriting-based communication for a wide range of users with motor impairments.

## Results

We surgically implanted two Utah arrays into the left motor cortex of a patient, specifically targeting the region surrounding the hand ‘knob’ area (inset, Fig. 1A). The patient, a right-handed individual in his 70’s, had experienced a C4-level spinal cord injury resulting in total sensory and motor loss below the shoulders. During experimental sessions, the patient was instructed to attempt to handwrite characters with his right-hand using chalk on a blackboard, following a video displayed on a screen (Fig. 1A). The video presented handwriting sequences of strokes and cohesions—representing the air connections between strokes—of a single character at an overall consistent speed (Fig. 1B). Fig. 1C illustrates the smoothed velocity profiles in the *x*- and *y*-direction during the writing of a character that comprised of three strokes and two cohesions, with each stroke or cohesion exhibiting a bell-shaped velocity curve.

**Fig. 1.**
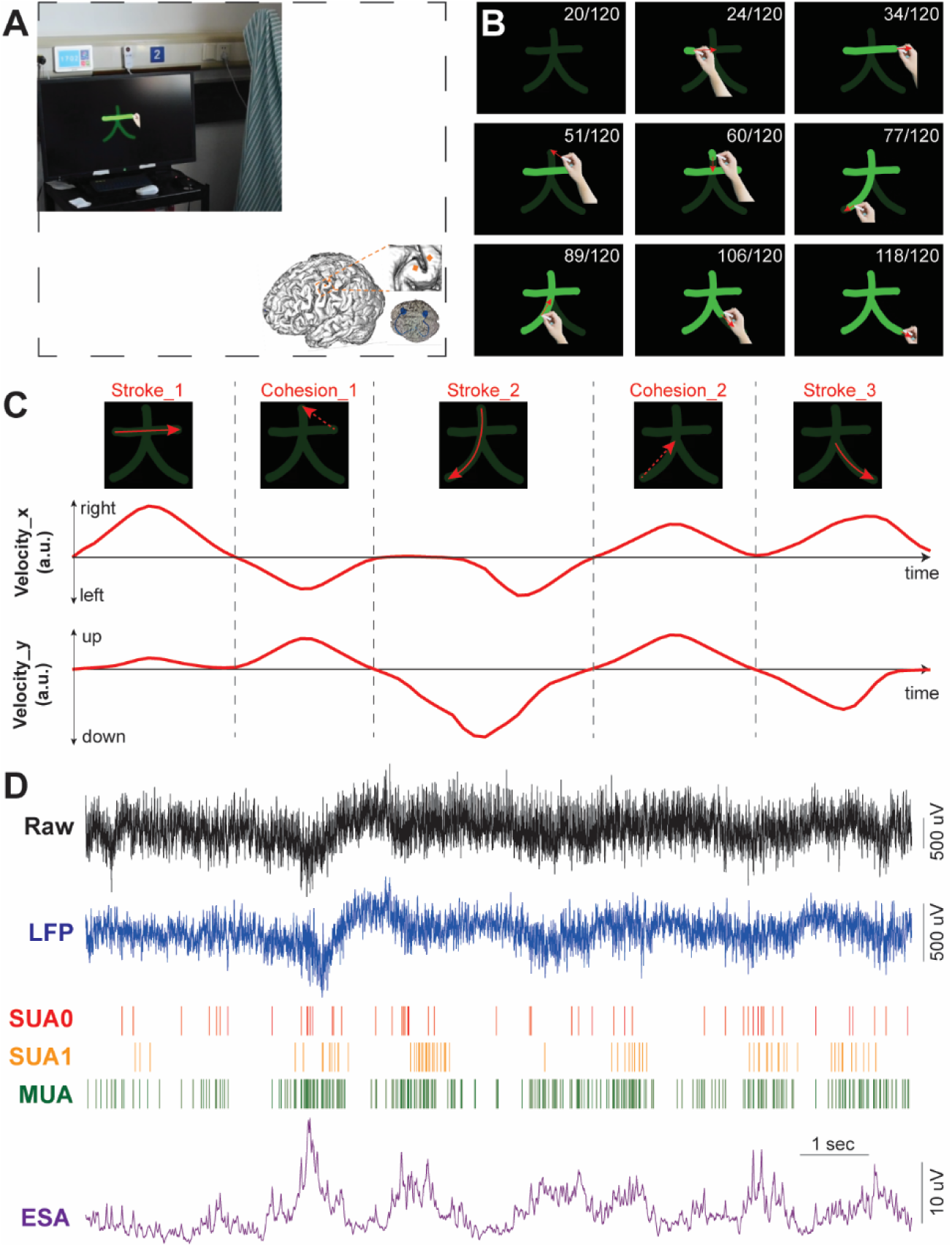
Experimental setup and neural signal recording. **(A)** The subject with recording cables connected was imaging handwriting with his right hand following the animation showed on the screen. Inset: illustration of implantation position for the two Utah array (orange) and two connectors (blue) in the left hemisphere. **(B)** Example frames of the animation video writing a Chinese character ‘大’ (big). The red arrow indicates the moving direction (not shown in the experiment). The number in the upper right indicates current frame/total frame number (not shown in the experiment). **(C)** Velocity profile (red lines) in x and y directions for the character ‘大’. The dash lines separate the three strokes and two cohesions, which are represented by red solid and dash arrows, respectively. **(D)** Example of neural signals when handwriting a character, including raw signal (30k sampling, 0.3-7500 Hz) and other processed signal features, like LFP, SUA, MUA and ESA.

We asked the subject to write 180 Chinese characters in 6 sessions (30 characters per session, 3 times/character). These characters are commonly used in daily life, with an average of 7.06 ± 2.78 strokes per character. We recorded raw neural signals during the imagined handwriting process. From these signals, we extracted features from both low and high-frequency bands (Fig. 1D), encompassing a range of measurements such as local field potential (LFP), single and multiple unit activity (SUA and MUA), entire spike activity (ESA), etc. (see Methods). We demonstrated that the neural activity patterns for each character were highly distinct; SVM classifier based on SUA achieved nearly perfect discrimination accuracy of 98.2% ± 2.29% among 30-character in each session. The correct rate was comparable with previous classification results of imaginary handwriting of 30 English letters and punctuations [7].

### MSE versus DILATE loss for decoding

We then tried to decode the trajectories of the handwriting using a machine learning approach. One possible problem with our experimental paradigm was that the patient may not have the exactly same speed profile as that of the guiding video, i.e., the neural activity and the label velocity profile were misaligned. This problem is commonly encountered in motor imagery experiments with subjects who had lost their motor ability. Meanwhile, the misalignment was not uniform across a single trial, making the problem more severe. The mean square error (MSE) loss employed previously overlooked this misalignment, which could train suboptimal decoders that affected the decoding performance. Therefore, we employed a recently introduced DIstortion Loss including shApe and TimE, referred as DILATE [15], to specifically address this prevalent problem in practical BCIs.

When the prompted trajectory (Fig. 2A) and imaginary trajectory (Fig. 2B) are not aligned in velocity but have the same shape, the MSE loss still calculates the loss value based on a one-to-one matching as shown in Fig. 2C. DILATE, on the other hand, was designed to stretch or compress time-series data point-by-point to make them as aligned as possible. It consists of shape loss and time loss. The goal of shape loss, which is essentially a soft DTW, is to find the optimal time alignment path that minimizes the overall distance between two time series (Fig. 2D, left). The soft DTW replaces the hard minimization operation in DTW by introducing a differentiable soft minimization operation, which makes the temporal alignment path smoother instead of enforcing a strict match between time points. The time loss, on the other hand, is to constrain that the optimal time alignment paths between prediction and goal do not deviate too much from the one-to-one matching alignment paths (Fig. 2D, right).

**Fig. 2.**
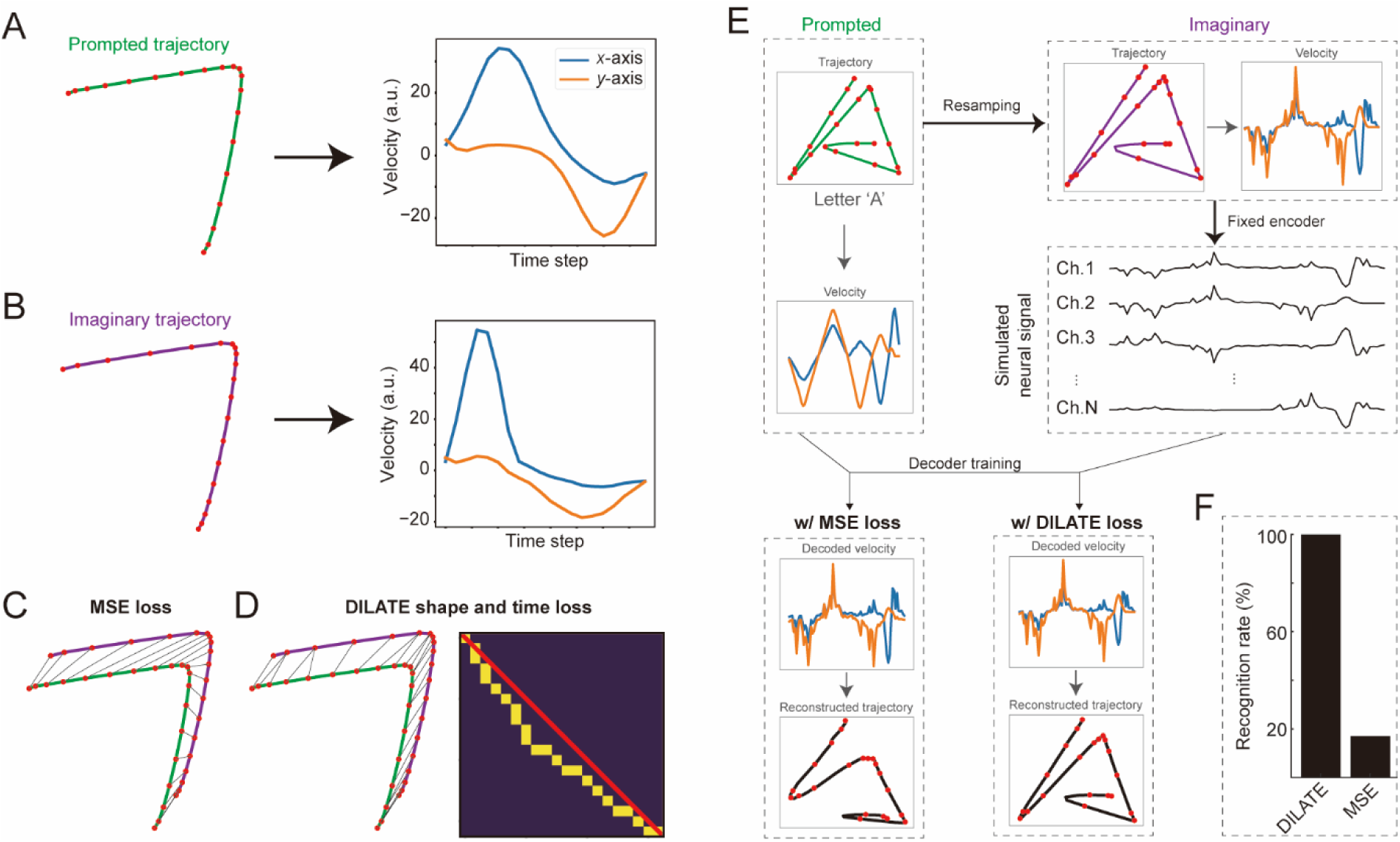
DILATE versus MSE loss for trajectory fitting. **(A)** Example of a prompted trajectory (dots indicate positions at equal time intervals) for handwriting imaginary and its corresponding velocity profiles in *x*- and *y*-axis. **(B)** Same as (A) but for an example imaginary trajectory actually adopted by the subject. Note the dots have different positions from (A) and thus velocity profiles are slightly different. **(C)** Schematic diagram for calculating MSE loss. The line between the prompted and imaginary trajectory indicates the point-to-point alignment of the MSE loss. **(D)** Schematic diagram for calculating DILATE loss. The line between the prompted and imaginary trajectory (left panel) and the sequence of yellow dots (right panel) indicates the shape and time alignment in DILATE loss. Shape loss calculates the distance between points, and time loss calculates the offset of the aligned paths with respect to one-by-one alignment (red line). **(E)** Schematic diagram of decoding simulation experiment. First, the prompted velocities were randomly resampled to obtain the imaginary velocities. Then, a fixed linear encoder is used to encode the imaginary velocities into simulated neural signals. Finally, based on the simulated neural signals and prompted velocities, the decoder is trained using MSE loss and DILATE loss respectively to obtain the decoded velocities to reconstruct the trajectories. **(F)** Recognition rate of the decoded trajectories (from letter ‘A’ to ‘Z’, 10 resamples per letter) in the simulated experiments using a generic handwritten character recognition software.

To demonstrate the advantage of the DILATE loss over the MSE loss when the signals are not aligned with the labels, we designed a simulated decoding experiment (Fig. 2E). In a database with handwriting trajectories of 26 English letters, we randomly resampled the velocity profiles for each letter under the condition that the reconstructed shape was guaranteed to be unchanged. Then a fixed linear encoder model transferred the resampled velocities into the simulated neural signals, which are, in principle, not aligned with the original velocities. We finally trained the same decoder using either MSE or DILATE loss and evaluated the reconstructed trajectories separately. The recognition rate using a generic handwriting recognition software for MSE decoding was 43.8%, whereas decoding with DILATE resulted in a recognition rate of 92.3% (Fig. 2F). We also did the same simulation with the180 Chinese characters used for the patient and the results showed recognition rates of 100% and 16.7% for DILATE and MSE loss, respectively. This comparison demonstrated that DILATE effectively addresses the misalignment issue, enabling the construction of accurate decoders.

To illustrate the reason why DILATE outperformed MSE under conditions of misalignment, we conducted a simulation in a simpler scenario (Fig. S1). Assuming that the imaginary velocity and the prompted velocity *y* were identical copies but only with a overall delay in time (Fig. S1A), and the neural signal *x* was identical (i.e., linear) to the imaginary velocity. We assumed a linear decoder (*y = ax + b*) to map the neural signal to the true velocity. When using MSE to train the decoder, the smallest loss value was found at *a* = 0 and *b* = 1, which was basically a flat line (Fig. S1B). In contrast, DILATE minimized at *a* = 1 and *b* = 0, which was an identical function, i.e., the shape was retained regardless of the delay (Fig. S1C). This result indicates that DILATE loss valued the overall shape matching (after distortion) instead of pursuing overall one-to-one error minimization as in MSE, which is exactly what we need in the handwriting BCIs where accurate path reconstruction is critical.

### Decoding and recognizing handwriting trajectory with MSE loss

We first demonstrated the decoding framework with conventional MSE loss. To realize brain-to-text translation, we fit the neural activities into the velocity of the handwriting, reconstructed the trajectory by performing an integration along the path, and then recognized the trajectories as standard texts using machine learning approaches (Fig. 3A). We trained both linear Kalman filter and nonlinear long short-term memory (LSTM) network using leave-one-character-out cross-validation for trajectory fitting. The goal of these two decoders was to minimize the MSE between the prompted writing velocity and the decoded velocity. The decoding results of various types of neural signals, quantified as correlation coefficient (CC), were presented in Fig. 3B. Across all scenarios, the LSTM demonstrated superior fitting outcomes compared to the Kalman filter. Notably, ESA yielded significantly better results than all other signal features, with an average CC of 0.753 ± 0.18. To provide a qualitative illustration of how the reconstructed trajectories varied with different CC values, we showcased five example reconstructions in Fig. 3C, with CC values ranging from 0.1 to 0.9. Generally, a reconstruction with a CC exceeding 0.5 would result in a human recognizable shape. However, quantitative results were required to assess the quality of reconstructed trajectories.

**Fig. 3.**
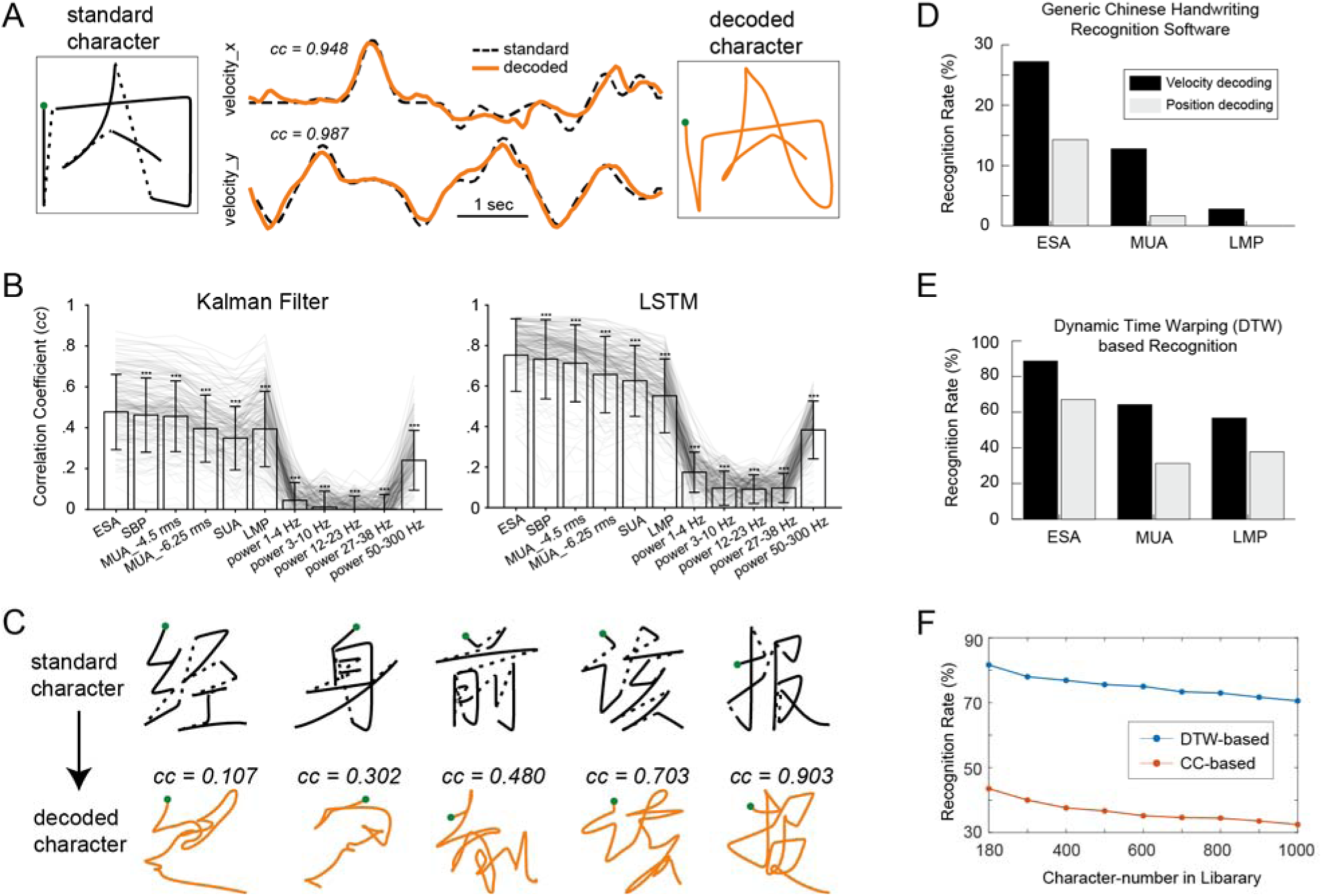
Handwriting trajectory fitting with MSE loss. **(A)** Example of a standard Chinese character ‘内’ (solid for strokes and dash line for cohesions, left panel) and the decoded trajectory (orange, right panel). The corresponding velocity profiles in x and y directions were showed in the middle panel. Green dots indicate the start of the handwriting. **(B)** The fitting correlation coefficients (CC) of the 180 characters for Kalman Filter (left panel) and LSTM (right panel) decoder with various kinds of neural signals. See text for the abbreviation. Asterisk (***) indicates significant difference between ESA and other signals (paired signed-rank test, *p* < 0.001). **(C)** Five more fitting examples (‘经’, ‘身’, ‘前’, ‘该’, ‘报’) with fitting CC ranging from 0.1 to 0.9. **(D)** Recognition rate of the 180 decoded characters using a generic handwriting character recognition API (from teshuzi.com). The decoding results for LMP, MUA, and ESA were illustrated. Decoding scheme for both velocity (black bar) and position (gray bar) was tested. **(E)** Same as (D) but for dynamic time warping (DTW) based recognition method which calculates similarity between the decoded trajectories and the standard Chinese character trajectories (180-character in library). **(F)** DTW- and CC-based recognition rate as a function of character number in the library.

To objectively assess whether the decoded trajectories could be recognized as legible text, we initially utilized a generic handwriting recognition software to discern the continuous trajectory for each character. In this scenario, we used ESA, SUA and LMP for decoding and compared both speed and position decoding schemes. ESA velocity decoding yielded the highest recognition rate; however, only around a quarter (27.6%) of the trajectories could be recognized as correct Chinese characters (Fig. 3D). This was not surprise because the decoded trajectory for each character was essentially a single continuous stroke, which significantly deviates from conventional stroke-by-stroke handwriting patterns that were used to train the generic handwriting recognition software.

To recognize the trajectories correctly, we devised an innovative method that aimed at finding out the standard character that has the most similar speed profile with the decoded trajectories. The underlying concept was that each character would generate a unique and distinctive speed profile identifier along the writing process. To that end, we first built a library that encompassed the speed profiles for writing the standard 180-character. Then each decoded trajectory was z-score normalized and matched with the most similar standard character using dynamic time warping (DTW) algorithm. ESA with velocity decoding achieved the highest recognition rate, and this time approximately 81.7% of the trajectories could be correctly recognized (Fig. 3E). Given that the speed profiles were highly unique for each character, only a slight decrease in recognition rate to 70.6% was observed when the library expanded to 1000 characters (Fig. 3F). Comparing to CC based method, DTW permits temporal sequences to exhibit certain degrees of delay or stretching along the time axis, thereby enabling a more precise capture of the inter-sequence similarity. This suggested that the recognition method was sufficiently robust for recognizing a large number of characters.

### Trajectory fitting of Chinese characters with DILATE loss

We next attempted to use the DILATE loss to train the decoder to fit the velocity of the handwriting of Chinese characters from the neural signal and reconstruct the trajectories. We first searched the two hyperparameter tradeoff α (0.1-0.9) and smoothing factor γ (0.0001-1) in the DILATE, and found that these two parameters had little effect on the decoding performance (Fig. S2); the final choice of parameters was α = 0.5 and γ = 0.001 throughout the study. As the example character showed in Fig.4A, the decoding result with DILATE (Fig. 4B) resembled more to the decoding target in both shape and stroke positions, and has a smaller DTW distance (a metric to measure similarity between two time series, especially for locally scaled but morphologically consistent time series, see Methods) to the target than that of MSE (Fig. 4C). More examples of reconstructed trajectories were shown in Fig. S3.

**Fig. 4.**
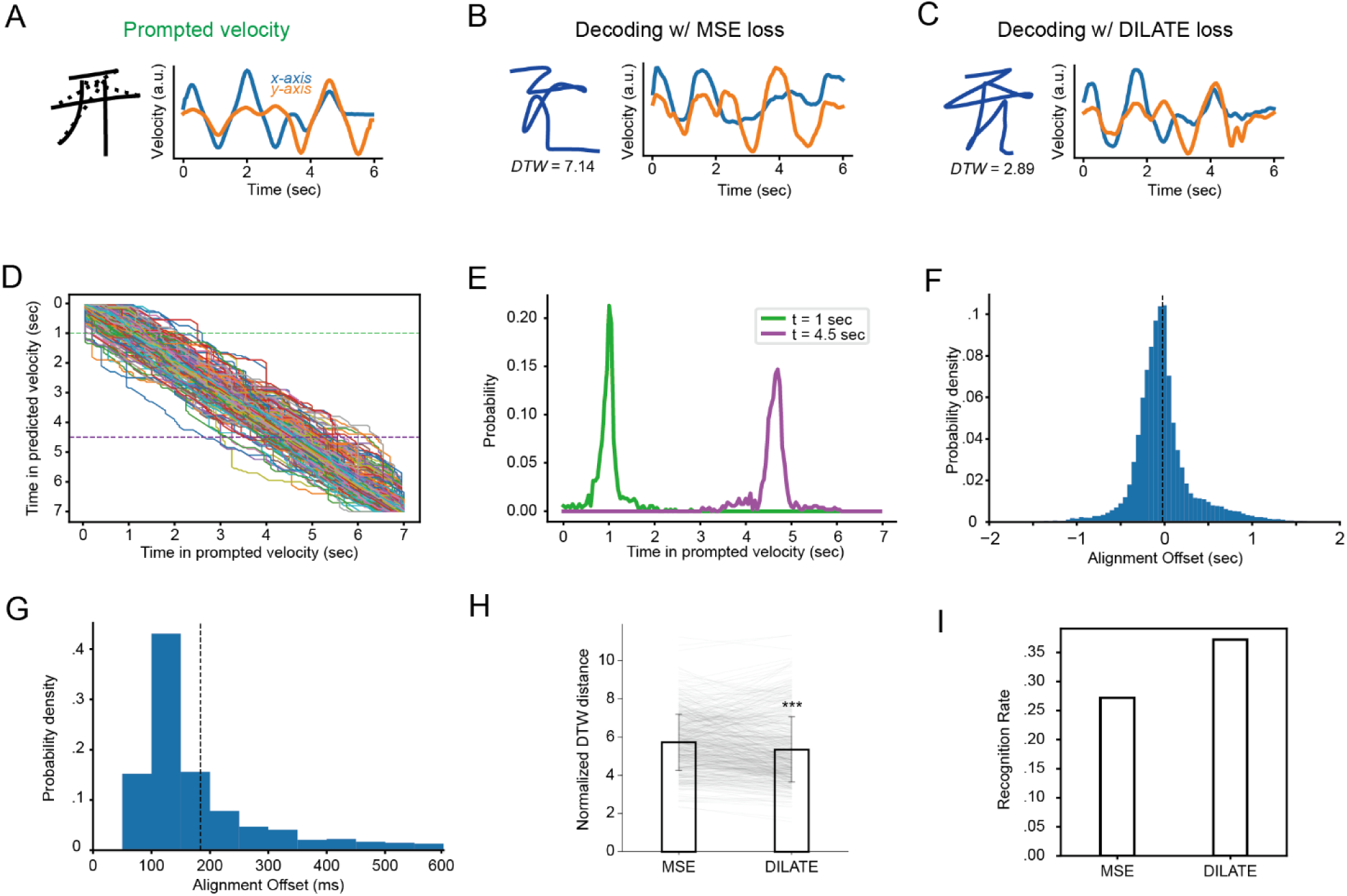
Trajectory fitting with DILATE loss. **(A)** Example of a standard Chinese character ‘开’ (solid for strokes and dash for cohesions, left panel) and the corresponding velocity profiles in *x*- and *y*-axis (right panel). **(B-C)** Same as (A) but for decoding result with MSE (B) and DILATE (C) loss. The DTW distances between decoded and prompted trajectories are indicated. **(D)** The alignment paths of DILATE for all the decoded and prompted velocities in all sessions. Each line represents one character in a trial. **(E)** Probability distribution of time steps that are aligned with the decoded velocity at time of 1 and 4.5 sec, i.e., two dash lines in (D). **(F)** Histogram of the alignment offsets for all trials. **(G)** Histogram of standard deviation of overall alignment offsets for predicted and label velocities for all trials. **(H)** Normalized DTW distance between all the decoded and prompted characters for MSE loss and DILATE loss (***, *p* < 0.001, paired signed-rank test). **(I)** Recognition rate of all the decoded trajectories using a generic handwritten character recognition software for MSE and DILATE decoding.

To reveal the actual alignment in DILATE, the normalized alignment paths in the algorithm for each trial (i.e., character) were plotted in Fig. 4D. The paths formed thick cloud along the diagonal, which indicated that the optimal alignment time was mostly time-distorted; if not distorted, the lines will be exactly diagonal. The probability density plots of at two slices at 1.0 and 4.5 sec (Fig. 4E) showed that the main alignment offsets were ranging from 500 to 1000 ms. To quantitatively measure the alignment offset, we calculated the single-point offsets for each character and plotted the histograms in Fig. 4F. The averaged alignment offset was -24.15 ± 356.07 ms, which indicated that the overall alignment offset is small but the local alignment offset fluctuates widely. We also calculated the standard deviation (SD) of the single-point offsets within each trial (Fig. 4G), revealing a wide SD of 183.62 ± 122.39 ms. These results indicates that the alignment in each trial was nonuniform and could be leading or lagging at different part of a trial.

Finally, we quantitatively compared the decoding results for DILATE versus MSE loss. The decoding results for all the 180 Chinese characters using DILATE were significantly better than those using MSE, with lower normalized DTW distance (5.35 ± 1.71 vs. 5.73 ± 1.47, Fig. 4H) and higher recognition rate (37.2% vs. 27.2%, Fig. 4I). Therefore, we demonstrated that the DILATE loss accounted for optimal alignment at each time step and achieved better decoding performance.

### Multi-day data fusion with DILATE loss

We then expanded the application of the DILATE loss to a multi-day scenario, where a more extensive dataset was created by integrating data from multiple days to train a new decoder, with the expectation of achieving improved performance (Fig. 5A). However, due to factors such as shifts in neural activity and data quality control, it was not guaranteed that the new decoder would perform better with an increased training dataset. We hypothesized that the DILATE loss, with its ability to effectively align neural activity with the target, could generate higher quality of training data. Consequently, we anticipated that the decoding performance derived from multi-day data could potentially outperform that of single-day data.

**Fig. 5.**
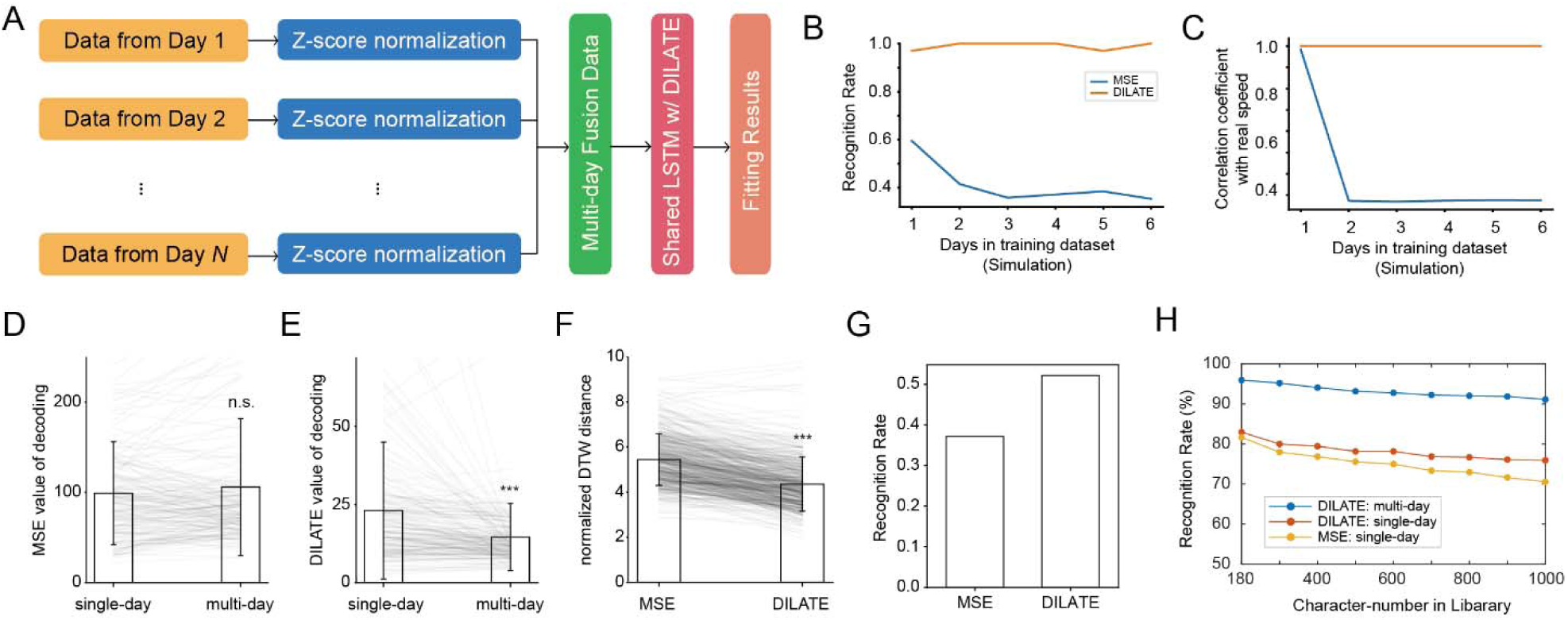
Multi-day data fusion with DILATE loss. **(A)** Schematic diagram of the multi-day fusion decoding framework. Multi-day neural data are mixed after z-score normalization and a shared LSTM network is then trained using the DILATE loss (or MSE loss) to obtain the fitting results. **(B-C)** The recognition rate (B), correlation coefficient (C) in multi-day simulation experiments (see text for details). **(D)** MSE loss values of single-day and multi-day decoding results with MSE loss (*n.s.,* no significance, *p* > 0.05, paired sign rank test). **(E)** DILATE loss values of single-day and multi-day decoding results with DILATE loss (***, *p* < 0.001, paired signed-rank test). **(F)** Normalized DTW distance for multi-day decoding results with MSE loss and DILATE loss (***, *p* < 0.001, paired signed-rank test). **(G)** Recognition rate for multi-day decoding results using a generic handwritten character recognition software for MSE and DILATE decoding. **(H)** DTW-based recognition rate as a function of character number in the library for DILATE (multi-day and single-day) and MSE (single-day) decoding.

To validate the hypothesis, we first conducted a simulation experiment akin to Fig. 3E, but with multi-day data. In the experiment, new data from up to 6 days were added to the decoding of characters in the first day. For each day, the velocities were subjected to random resampling to generate unaligned simulation signals. The DILATE decoding consistently sustained superior decoding performance, showing no sign of degradation as the number of days increased. In contrast, the MSE loss exhibited relatively inferior performance and eventually plateaued at a stable level (Fig. 5B). We then quantified the similarity between the decoded velocity profiles with the imaginary velocity (i.e., resampled ones), which represented the extent to which decoding speeds can still approach real imaginary speeds in the presence of label misalignment. For DILATE, the CC remained stable, whereas with the traditional MSE loss, there was a noticeable decline in similarity as the data in training set increased.

To evaluate DILATE on real-word data, we normalized the neural signals using z-score for each channel and combined data from all 6 days (30 characters/day) for training a shared decoder (Fig. 5A). In other words, each character now was decoded by decoders trained with 179 other characters, instead of 29 characters in single-day case. When trained with MSE loss, the average MSE loss values of the multiday decoding result were not significantly different from those of single-day decoding (105.83 ± 75.85 vs. 99.03 ± 56.99, *p* > 0.05, Fig. 5D). However, when trained with DILATE loss, the average DILATE loss values for the multiday decoding were significantly reduced compared to single-day decoding (15.17 ± 11.14 vs. 23.99 ± 22.76, *p* < 0.001, Fig. 5E). We then employed the normalized DTW distance as a standard metric to assess the multi-day fusion decoding outcomes with both loss functions. The decoding results achieved with DILATE were significantly superior than those of MSE (4.35 ± 1.20 vs. 5.44 ± 1.15, *p* < 0.001, Fig. 5F). Most notably, the multi-day fusion decoding using the generic handwriting recognition software yielded a recognition rate of 52.2%, substantially higher than that of the single-day decoding of 37.2% (Fig. 5G). When the DTW-based template matching method used, the recognition rate for DILATE multi-day decoding achieved 91.1% in a 1000-character database, far higher than DILATE and MSE single-day decoding. These results indicated that the DILATE loss effectively mitigated the challenges associated with multi-day data fusion problem in BCIs.

### Trajectory fitting of English letters with DILATE loss

To further evaluate the capabilities of DILATE-based decoding, we utilized a previously published dataset of imagined English letter handwriting [7]. In this dataset, intracortical neural activity was recorded from motor cortex of a paralyzed subject who attempted to handwrite letters and symbols following a cue within a fixed interval (Fig. 6A). The main difference with our study was that there was absence of video guidance for letter writing, which could result in more pronounced misalignment between the neural signals and handwriting movements. In their original study [7], the trajectory was reconstructed only with the fine aligned, trial-averaged neural activity.

**Fig. 6.**
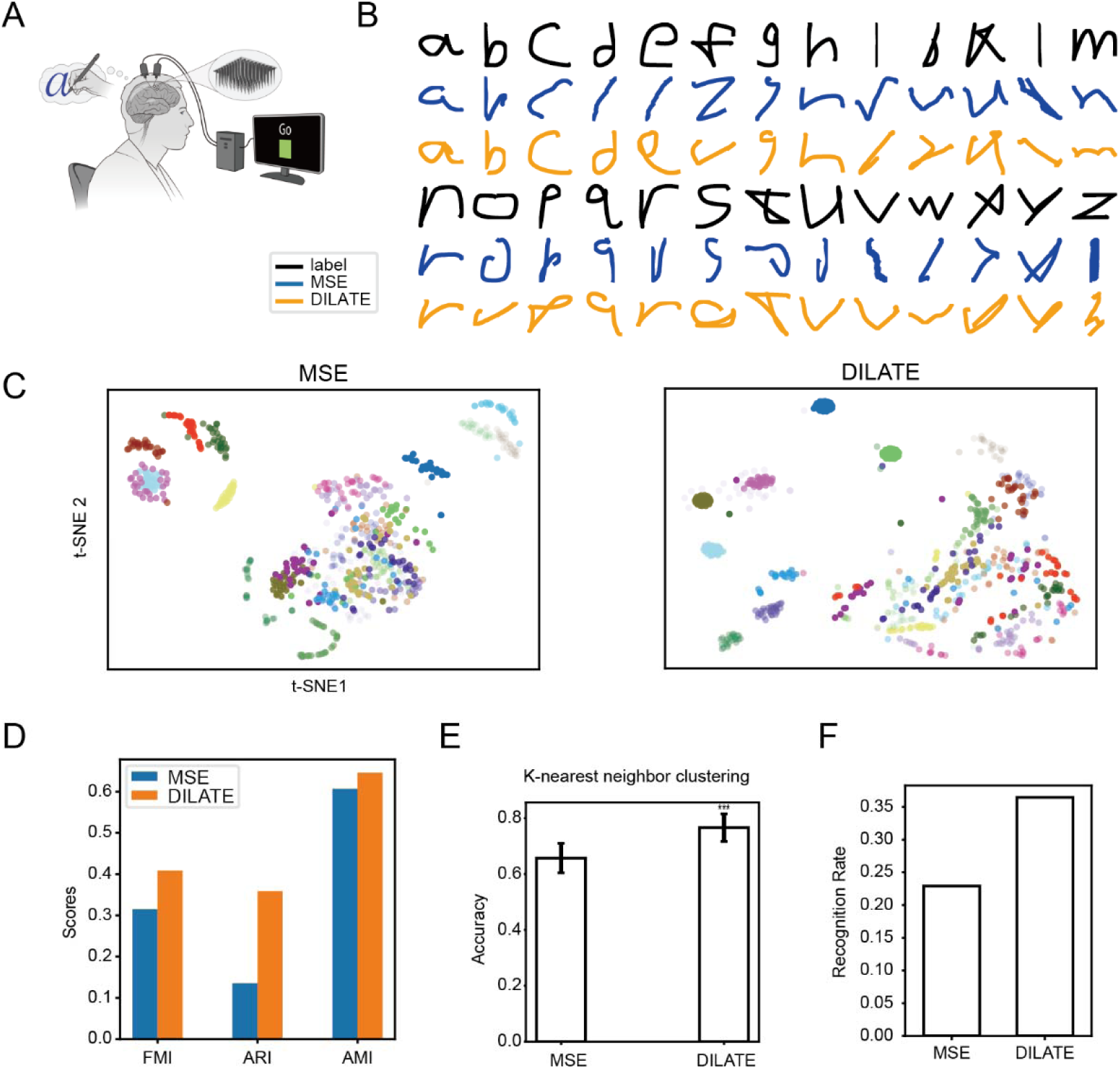
Trajectory fitting of English letters with DILATE loss. **(A)** Schematic of the imaginary English letters handwriting experiments (from Willett et al., 2021). **(B)** Example trajectories of all the letters reconstructed with MSE and DILATE loss. **(C)** Visualization of t-SNE dimensionality reduction for the decoding results with MSE loss (left panel) and DILATE loss (right panel). **(D)** Evaluation of the clustering with three metrics, FMI, ARI, and AMI, for MSE and DILATE decoding. **(E)** Classification accuracy with k-nearest neighbor for MSE and DILATE decoding (***, *p* < 0.001, t-test). **(F)** Recognition rate of the decoded letters using a generic handwritten character recognition software for MSE and DILATE decoding.

We used DILATE to investigate whether the neural signal could be automatically aligned at single-trial level, thereby improving the reconstructed trajectory. Fig. 6B presents the example label trajectories for the letters that were required to copy for the patient, along with the decoded trajectories using the MSE and DILATE loss, respectively. In general, the decoding results using DILATE outperformed those of MSE, yielding more human-recognizable letters.

We next quantitatively analyzed the decoding results. We first assessed the discriminability of the decoded results for both loss functions by applying t-SNE downscaling followed by HDBSCAN clustering (Fig. 6C). The decoding results using DILATE achieved better results in all three clustering evaluation metrics, including Fowlkes-Mallows Index (FMI), Adjusted Rand index (ARI), and Adjusted Mutual Information (AMI) (Fig. 6D). We then employed the k-nearest neighbor classification algorithm to categorize the decoding results for both loss functions. The classification results using DILATE achieved a correct rate of 76.61 ± 4.97%, which was significantly better than the 65.71 ± 5.25% obtained with MSE (Fig. 6E). Finally, we evaluated the decoding results with the two loss functions by direct feeding the decoded trajectories to the generic handwriting recognition software. The decoding results using DILATE achieved a recognition rate of 36.47%, outperforming the 22.93% achieved with MSE (Fig. 6F). These results demonstrated superior decoding performance of DILATE over MSE loss on both Chinese character and English letters handwriting dataset.

## Discussion

In this study, we recorded intracortical neural activity from a human patient during video-guided imaginary handwriting and reconstructed trajectories of the imagined handwriting using a novel decoding framework that incorporates DILATE loss. The findings revealed that DILATE outperformed traditional MSE loss across both simulated and experimental datasets, including both single-day and multi-day decoding scenarios. More than 52% of the decoded trajectories were recognized as legible texts using generic handwriting recognition software; employing DTW-based template matching methods, the recognition rate achieved up to 91.1% within a 1000-character database. This innovative decoding framework was further applied to a previous imaginary English letter handwriting dataset, which had been solely used for classification purpose, and demonstrated the ability of single-trial trajectory reconstruction and better letter-recognition with DILATE loss. By addressing the misalignment issue with DILATE loss in clinical BCIs, our results enabled high-performance trajectory-based brain-to-text translation, which hold promise for applications across universal languages. This approach thus opens new avenues for individuals with motor deficit to communicate through written languages in any form.

Our study introduced the DILATE loss [15] for the first time in the clinical BCIs, which optimally aligned neural signals and handwriting labels during the training of the regression model, and achieved superior decoding performance in both Chinese character and English letter handwriting dataset. Label misalignment with ‘thoughts of motion’ is a prevalent issue for training an accurate decoder in motor imagery based BCI, especially when continuous output is required. As a contrast, conventional-used MSE loss performs poorly with signal distortions conditions [15-17, 21], thus not feasible for this clinical BCI application. Alternatively, previous studies have attempted to solve the signal misalignment problem with time-warping method [7, 22, 23], which primarily align signals from different trials under identical conditions. However, this method does not ensure that the aligned signals correspond accurately with their kinematic labels, which is the required in our application. Another speech BCI [6] introduced connectionist temporal classification (CTC) loss [24] to alleviate the problem of lack of ground truth, but this approach is limited to sequence alignment for classification tasks and cannot be extended to regression task in this study. Our results thus provide an optimal way to address the misalignment challenge in the regression-based motor imagery experiments, eliminating the need to pre-align the signal and label before decoder training. This strategy is applicable to decoder training across various motor imagery experiments.

We introduced the trajectory-based handwriting paradigm for BCI, i.e., decoding the trajectory of imaginary handwriting and then recognizing the trajectories as standard texts. Previous study showcased the trajectories of single-letter handwritings only with trial-averaged neural activity [7]. Due to the precise alignment between neural activity and handwriting kinematics in our study, we have been able to reconstruct complex writing trajectories as human recognizable characters on a single-trial basis. Other researches have predominantly concentrated on decoding straight movements in arm-reach distances to control computer cursors [3, 25] or prosthetics [2, 26]. Preliminary studies have also explored the decoding of simple curved drawings in monkeys [18, 27]. One critical challenge was the extraction of accurate speed profile which is nonlinearly encoded [28]. Our study demonstrated the ability to reconstruct the intricate handwriting trajectory, which occurs within a significantly smaller range but encompasses distinct spatial and temporal dynamics [29]. Meanwhile, we recognized the decoded trajectories using both generic handwriting recognition software and custom template-matching methods. Handwriting recognition and Optical Character Recognition (OCR) have reached a high level of sophistication and are widely utilized in contemporary applications [30, 31]. However, these standard recognition techniques are not well-suited for recognizing trajectories in our study, which are basically one-touch-writing trajectories that are distinct from normal writing patterns. This is one of the reasons that the recognition rate using this method is relatively low (<52%) in our study. On the other hand, leveraging the fact that each character has its own unique speed profile, the template-matching method we developed achieved much higher accuracy and is robust to the size of database. To the best of our knowledge, this research marks the inaugural attempt to reconstruct complex handwriting movements for brain-to-text communication. This novel strategy extends the application of handwriting BCIs to encompass any written language, be it Latin-based or non-Latin, as it enables the decoding of any written trajectory as it is, thereby broadening the horizons for individuals seeking enhanced communication capabilities.

The DILATE loss were also demonstrated to improve decoding performance by multi-day data fusion. The fusion of multi-day data to enhance the performance of decoder has been a difficult problem in BCI due to the two opposite effects. On one hand, fusion of multi-day data increases the amount of data to train the decoder, which could potentially enhance the performance. On the other hand, if the data are inhomogeneous, due to instability of neuronal recording or low data quality, the performance of decoder could be deteriorated. Most of the past studies align the multi-day data by mapping or extracting the low-dimensional latent dynamics using methods like canonical correlation analysis [32], non-negative matrix factorization [33] or linear mapping [7]. We argue that if we could align the single-day data more accurately to enhance the data quality, the multi-day would be solved automatically. Our results showed that the DILATE loss effectively reduced the misalignment problem and further enhanced the decoding performance comparing with the single-day results. For MSE loss, the two opposite effects on decoding performance canceled with each other and achieved not significant results. Thus, we developed a novel methodology with DILATE to cope with multi-day fusion problem in imagery-based BCIs.

Our study, while illuminating, has several limitations that warrant acknowledgment. First, we did not conduct online decoding experiments to verify the effectiveness of the DILATE loss in an online experimental scenario. Nonetheless, this approach remains a valuable starting point for constructing an initial high-performance decoder, which can be further refined during online testing. Second, our study still considered handwriting as a 2D plane movement, rather than employing a 3D or multi-dimensional model. Future research should integrate these additional dimensions to fully account for the variations observed in neural data, and potentially enhance the decoding performance further with these additional insights.

Through training decoders with optimal-aligned signals, our findings could be directly applied in clinical BCIs to enhance the decoding performance. This advancement will expedite the translation of BCI researches into practical human applications, broadening the scope of BCIs with handwriting or even drawing-related paradigms.

## Materials and Methods

### Participant and surgery

The participant enrolled in this study was a right-handed individual, who had experienced a C4-level spinal cord injury and resulted in total sensory and motor loss below the shoulders. The microelectrode implantation surgery was conducted about 3 years after the injury in his 70’s and data collection for this study was at around 2.5 years after the surgery [34]. All clinical and experimental procedures received approval from the Medical Ethics Committee of the Second Affiliated Hospital of Zhejiang University and were registered in the Chinese Clinical Trial Registry (chictr.org.cn; registration number: ChiCTR2100050705).

Two 96-channel Utah microelectrode arrays (Blackrock Microsystem, USA) were implanted into the left precentral gyrus, specifically targeting the hand ‘knob’ area of motor cortex (Fig. 1A inset). The location of implantation was identified using functional magnetic resonant imaging (fMRI) prior to surgery when the participant imaging reaching and grasping movement.

### Video-guided handwriting paradigm

To guide the motor imaginary process for the patient, a handwriting video was played on the computer monitor. The video consisted of stroke-by-stroke writing animation of a specific character, leading by a hand with chalk (Fig. 1A). The patient was asked to attempt to write the same character with chalk on a blackboard following the guidance. We also asked the patient to write on a paper with pen, basically the classification results were similar. We kept using chalk on blackboard paradigm based on the patient’s preference. A typical trial started by showing the character (in dark green) on the screen (500 ms) followed by an auditory prompt of the character’s pronunciation (1000 ms). After a short delay (300 ms), a sound cue was issued and the writing animation started. The writing consisted of both strokes and cohesions, i.e., air connection between strokes. The written strokes were highlighted as light green and the cohesions were simplified as a direct line between the end of current stroke and the start of next stroke. The duration of writing depended on the length of the character, ranging from 4 to 8 seconds, which is a little bit longer than normal writing speed to adapt to the patient. The speed for each character and cohesion was constant, i.e., the duration of each stroke or cohesion is proportional to their lengths.

The handwriting videos were artificially synthetic. Firstly, the sequences of two-dimensional coordinate for writing each character were extracted from standard font of that character using ‘GetData Graph Digitizer’ software. Secondly, each segment, defined as a straight line or an approximation of a straight line before sharp inflections, was labeled as stroke or cohesion and converted the coordinates into velocity sequences. The duration for each segment was proportional to the ratio of the segment length to the total length, and the velocity profiles in *x*- and *y*-direction were defined as:

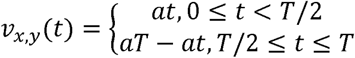

Where *T* represents total duration of that segment, and *a* is the scaling factor to fit the duration of the segment. Lastly, the handwriting animations were created frame-by-frame according to the velocity profile above using MATLAB. The video had a black background, and there was a static dark trace of the entire character before the actual writing starts. The strokes are represented by light green lines with thick width over the static dark characters (Fig. 1B). The position and velocity data used for decoding were 5-point smoothed version of the actual traces (sampled at 20 Hz), which resembled a bell-shaped profile (Fig. 1C).

### Data collection sessions

Neural data were recorded when the subject attempted to write various characters during 1-2 hours sessions on scheduled days. During the experimental sessions, the patient was seated in a wheelchair with hands resting on a table. A computer monitor was setup in front of the patient for task visualization. Two cables were connected from the patient’s head connectors to the NeuroPort data acquisition system (Blackrock Microsystem, USA), which recorded both neural signals and task timings (through serial port) simultaneously. The character dataset used in this study included 270 complex Chinese characters, 180 of which were recorded with raw data and various signal features could be extracted and used for decoding analysis (Fig. 3). For each character, 2 blocks and 3 repeats/block were conducted per session.

### Neural signal preprocessing

Neural signals from each channel were amplified, filtered (0.3-7500 Hz) and digitized at a sample rate of 30 kHz using NeuroPort. Various signal features were then extracted, including:

1. Single-unit activity (SUA), which was extracted online after further filtering (250-5000 Hz) with a threshold of -6.25 times root mean square (rms). Single units were isolated offline using Offline Sorter (Plexon, USA).
2. Multi-unit activity (MUA), which was extracted offline from the further filtered data (250-5000 Hz) using different threshold at -4.5 and -6.25 rms. No further spike sorting was applied.
3. Local filed potential (LFP), which was obtained by low-passing (below 500 Hz) of raw signal and down sampled to 2000 Hz. To reduce sporadic outliers, extremes exceeding ±3 times the standard deviation from mean were clipped, followed by a third-order Butterworth lowpass filter. Then the mean powers for each frequency band (1-4, 3-10, 12-23, 27-38, 50-300 Hz) were calculated as signal features.
4. Local motor potential (LMP), which was the moving averaged of LFP in non-overlapping 50 ms windows [35].
5. Entire spiking activity (ESA), which was obtained by applying a first-order Butterworth high-pass filter (300 Hz) on raw signal, rectifying by taking the absolute value, first-order Butterworth low-pass filtering (12 Hz), and finally down sampling to 1 kHz [13].
6. Spiking-band power (SBP), which was obtained by applying a second-order Butterworth bandpass filter (300-1000 Hz) to the raw signal, rectifying by taking its absolute value, and finally down sampled to 2 kHz [36].
7. Continuous multiunit activity (cMUA), which was obtained by applying third-order Butterworth bandpass filtering (300-6000 Hz) to raw signal, squared, low-pass filtering using a third-order Butterworth filter (100 Hz), clipping negative values, square rooted, and finally down sampled to 1 kHz [37]. We have found cMUA had high correlation coefficient (above 0.87) and similar decoding results with ESA, thus was not used for further analysis.

To identify the actual timing of imaginary handwriting after animation start, we performed principal component analysis (PCA) and found a significant change of neural activity in PC1 and PC2 occurred at around 300 ms after the cue. Subsequent decoding analysis confirmed that a delay of 300 ms achieved the best results. Therefore, we aligned the writing kinematics with the 300-ms-shifted neural in all following analysis. The bin size to average the neural activities were also tested in a classification decoding task, ranging from 50 to 400 ms, and confirming that a bin size of around 200 ms yielded the best results. Thus, all the neural signal features above were binned with overlapping 200 ms window and shifted 300 ms to align with handwriting kinematics.

### MSE-based trajectory fitting and recognition

To fit the trajectory of the imagined handwriting movement from the neural signal features, we utilized both Kalman Filter (KF) and long short-term memory (LSTM) as decoder [13]. The KF uses linear system state equation and the input-output data observed to estimate the system’s state optimally. The KF employs a recursive approach for state prediction and state updates as follows:

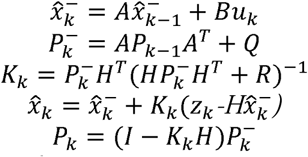

where 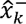 is the predicted state value, 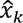 is the optimal estimate of the state, *A* is the state transition matrix, *B* is the control input matrix, *H* is the state observation matrix, *Q* and *R* represent the covariances, which respectively characterize the deviations of the state values and observation values.

The LSTM is a type of recurrent neural network (RNN) designed specifically to solve the issue of long-term dependencies in traditional RNNs. The core of LSTM is the cell state, which serves to stably preserve long-term memory in the model. LSTM utilizes gate mechanisms to control the removal or addition of information to the cell state. The forget gate determines which information should be discarded from the cell state, the input gate determines which new information should be added to the cell state, and the output gate determines the features of the cell state to be outputted. The description is as follows:

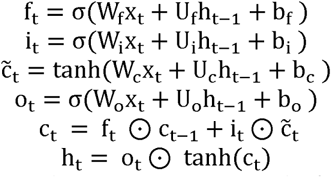

where *x* represents the input, *h* represents the output, *f* represents the forget gate, *i* represents the input gate, *o* represents the output gate, *c* represents the cell memory. The symbols *σ* and ⊙ represent the sigmoid activation function and element-wise multiplication operator. The number of units in the LSTM was 512 and the network was trained with batch size of 1, dropout rate of 0 and learning rate of 0.001.

The fitting models were cross-validated using a leaving-one-character-out method, in which, all the repeats for the same character to be tested was excluded for training the model. Both velocity and position of the handwriting were used to decode the trajectory of characters. For velocity model, an additional step that integrating velocity along the path was calculated to reconstruct the position, i.e., trajectory. Finally, we used Mean Squared Error (MSE) and Pearson’s Correlation Coefficient (CC) as evaluation metrics for decoding performance and paired Wilcoxon signed-rank tests to assess statistical differences in decoding performance between different features and decoding methods.

Decoded handwriting trajectories were recognized as text in two ways. Firstly, the trajectory for each character was fed into an online generic handwriting recognition software through their APIs (teshuzi.com). The first Chinese character output by the algorithm, which has the highest similarity score, was selected as the recognition outcome. Secondly, we recognized the decoded trajectories by matching them against a database of velocity profiles from standard characters. To accomplish this, we extracted trajectories for up to 1000 commonly used characters (using methods above) and converted them into their corresponding velocity profiles. The 180 characters tested in this study were part of this library, but the velocity profiles in the library were not identical with the velocity prompted to the subject (due to different sampling). Dynamic Time Warping (DTW) and correlation coefficient (CC) were employed to quantify the similarity between the decoded velocity and velocity profiles in the library. DTW permits temporal stretch and delay, thereby enabling a more precise capture of the inter-sequence similarity. However, the computation load was high for DTW and the fastDTW algorithm was employed to compute the DTW distances. The character with the highest similarity score, as determined by DTW or CC, was selected as the final recognition result.

### DILATE loss and simulation

The DILATE loss is based on smooth appropriation of DTW with a temporal term [15]. For two *d*-dimensional time series *y* and *z* of length *m* and *n*, the DTW finds an optimal alignment path *A* ⊂ {0, 1}*^m×n^*, where *A_i, j_* = 1 when *y_i_*and *z_j_* are aligned. In the set of admissible alignment paths *A_m, n_*, the alignment paths are always connected to (*m*, *n*) from (1, 1) and when *A_i, j_* = 1, the next alignment path point can only be *A_i+1, j_*, *A_i, j+1_* or *A_i+1, j+1_*. Δ(*y*, *z*) is the point-to-point loss matrix for the sequences *y* and *z*, where the point-to-point loss is computed using the Euclidean distance, i.e., 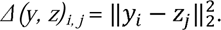 The DTW computes the alignment path with the smallest cumulative cost, that is

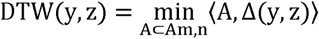

where ⦑·,·⦒ denotes the inner product of two matrices.

To deal with the non-differentiability of DTW, the shape loss part of DILATE introduces a differentiable minimization function that replaces the hard minimization operation in DTW, so that the loss function gains global differentiability. The soft minimum operation and shape loss are as follows:

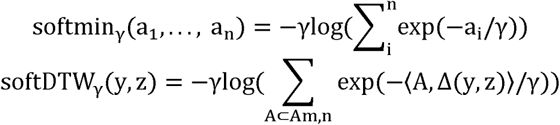

To make the model not only focus on the similarity of sequence shapes when predicting, but also consider the time accuracy of the prediction results, DILATE adds a time loss function along with softDTW. This function is an improved version of the Temporal Distortion Index (TDI), which is a common time similarity. The TDI is defined as

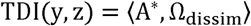

where A* = argmin_A⊂Am,n_⦑A,Δ(y, z)⦒ is the DTW optimal path, Ω_dissim_ is a matrix of size *m*×*n*, where Ω_dissim_(i,j) = (i - j)^2^, serves to penalize alignment point pairs that deviate from the diagonal of the path matrix.

The original version of TDI is non-differentiable because the optimal path A is non-differentiable with respect to Δ. To deal with this problem, the time-loss part of DILATE handles this using a relaxed optimal path A_γ_^*^, which is defined as the gradient of the softDTW_γ_. The relaxed optimal path A_γ_^*^ and time loss are as follows:

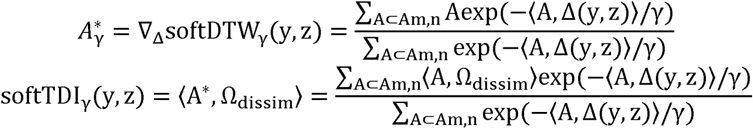

Ultimately, the DILATE loss is weighted sum of the shape loss and the time loss, that is

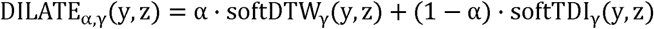

To test the performance advantage of using DILATE loss over MSE loss, we designed a simulation experiment with the following steps:

1. Reconstruct the prompted velocity sequence *V* into a trajectory sequence *P* and assume that the velocity between two points of the trajectory sequence is uniform.
2. Randomly sample time points from a uniform distribution and sort them, with values ranging from 0 to the time step of the trajectory sequence, and the number of values is the time step of the trajectory sequence, calculate the positions of the corresponding time points in the trajectory sequence to obtain the resampled trajectory sequence *P’*, and differentiate to obtain the simulated imaginary velocity sequence *V’*. The resampling time points are random and different for different simulated trials.
3. A fixed linear layer neural network is used as an encoder with input *V’* to generate the simulated neural signal *S’*.
4. Train the linear layer neural network with a 5-fold cross-validation scheme using two losses separately, with *S’* as the signal and *V* as the target, to obtain the prediction *Vp*.
5. Test the errors of *Vp* and *V’* using MSE, CC metrics, and test the direct recognition rate of *Vp*.

To visualize the applicable scenarios and advantages of DILATE, we simplify the simulation experiments. We use one-dimensional speed for simulation, and assume that the prompted speed and imaginary speed are as in Figure S1A. since both encoder and decoder are linear layer neural networks, in fact, *Vp* = *a*\**V’* + *b*, where *a* and *b* are the weight and bias. Two kinds of loss values are computed for different *a* and *b*. Then we plotted the 3-dimensional loss topography (as shown in Fig. S1B-C) separately and calculated the minimum value points. Comparing the difference between the two loss minima points, the advantage of DILATE loss in data and label misalignment scenarios can be obtained.

### DILATE-based trajectory fitting

To assess the efficacy of the DILATE loss in fitting handwriting trajectories, we employed an LSTM decoder configured identically to one utilizing MSE loss, except that the loss function was replaced with DILATE. We first z-score normalized the model output and the labels by dimension, and then calculated the DILATE loss values. This reduces the impact of the difference between the initial model output order of magnitude and the target on the loss calculation. The models were validated using a leave-one-character-out cross validation approach as above. Additionally, we explored the impact of the two hyperparameters— tradeoff α (ranging from 0.1 to 0.9) and smoothing γ (ranging from 0.0001 to 1)—in DILATE on decoding outcomes with one example session. We adopted normalized DTW as a quantitative metric to evaluate the decoding performance for both loss functions. Alongside this, we incorporated the previously mentioned online handwriting recognition software to objectively assess the recognition rate for trajectories decoded by the two loss functions.

Furthermore, we quantified temporal distortions of DILATE’s decoding outcomes relative to the labels, by calculating DTW to ascertain optimal alignment paths A* between decoding results and labels, where A^*^ = argmin_A⊂Am,n_(A,Δ(y, z)). Then the offsets for each point across all trials were concatenated and the standard deviation (SD) of these offsets (after absolution) within each trial were calculated.

### Multi-day data confusion

We extended the simulation experiment above for multi-day data fusion decoding. The amount of data in the training set were increased without changing the test set to simulate an increase in the amount from multi-day. The increased training data were the velocities obtained from new character trajectories after random resampling, and the simulated neural signals were obtained after the same fixed encoder. The encoder for all characters was the same, but the resampling is random for each character. Finally, the decoding performance with increasing training data volume was tested. We employed MSE, CC and recognition rate as evaluation metrics for the simulation experiments.

To perform multi-day data fusion decoding in real dataset, we first applied z-score normalization of each day’s neural signals by channel, and then concatenated them into a single dataset for training the decoder, with either MSE or DILATE loss function. The decoded velocities were subjected to calculate the MSE and DILATED loss respectively to evaluate the single-day versus multi-day decoding. Furthermore, normalized DTW, which depicts the resemblance between the decoded trajectory and the label, was calculated to evaluate the quality of character reconstruction for both loss functions. We used paired Wilcoxon signed-rank tests to assess statistical differences in decoding performance.

### Decoding trajectory for English letters

The English handwriting dataset was from the previous study [7]. In brief, the subject, with two microelectrode arrays implanted in the motor cortex, was instructed to attempt to handwrite 26 lowercase English letters following a ‘go’ cue. The subject finished one single letter writing within a fixed interval without any video guidance. The MUA activity was recorded through the two 96 electrode intracortical arrays. The dataset also provided the label trajectory of each letter, from which we extracted the speed profiles for decoding. The speed profiles were linearly interpolated to the same length as the neural activity. Fitting experiments were then performed using the same LSTM model as above with the two loss functions. The fitting models were cross-validated using a leaving-one-letter-out method.

We first downscaled the fitted results to 2 dimensions using t-distributed stochastic neighbor embedding (t-SNE), then clustered them using the HDBSCAN method [38], and finally evaluated the clustering results using metrics of Fowlkes-Mallows Index (FMI), Adjusted Rand index (ARI), and Adjusted Mutual Information (AMI). Next, we randomly split the fitting results into 90% training set and 10% test set and classified them using k-nearest neighbor (KNN) classifier. We repeated the classification experiment 100 times and statistically compared the classification accuracy (t-test). Finally, we compared the fitting performance of the two loss functions using the generic recognition software.

## Data Availability

All data produced in the present study are available upon reasonable request to the authors

## Acknowledgments

This work was supported by STI 2030—Major Projects (2021ZD0200404), National Natural Science Foundation of China (62336007), Pioneer R&D Program of Zhejiang (2024C03001), the Starry Night Science Fund of Zhejiang University Shanghai Institute for Advanced Study (SN-ZJU-SIAS-002), and the Fundamental Research Funds for the Central Universities (2023ZFJH01-01, 2024ZFJH01-01). The authors thank Mr. Xiang Li for software development.

## Author contributions

Conceptualization: GX, YW, YH; Methodology: GX, ZW, KX, JZhu, YW, YH; Investigation: GX, ZW, JZhu, YH; Visualization: GX, YH; Supervision: JZhang, YW, YH; Writing—original draft: GX, YH; Writing—review & editing: KX, JZhang, YW, YH.

## Competing interests

Authors declare that they have no competing interests.

## Data and materials availability

All data in the main text or the supplementary materials are available upon request.

## Figures and Tables

**Supplementary Fig. 1.**
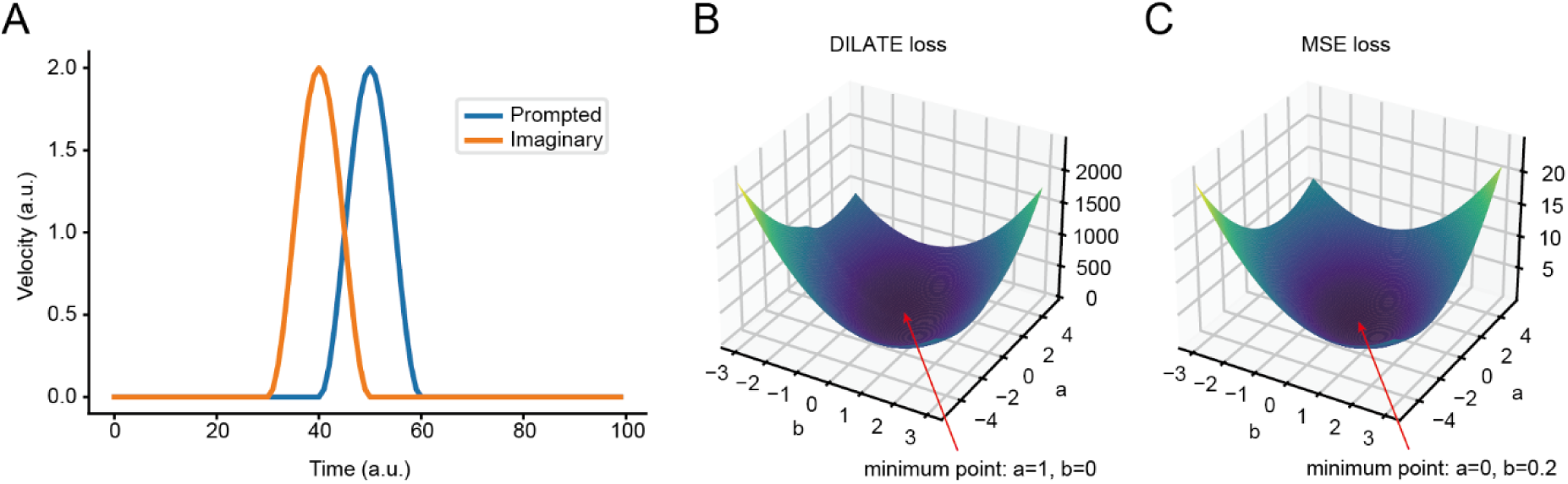
DILATE loss handles unaligned data better than MSE loss. (**A**) Schematic representation of the imaginary and prompted speeds. The peak of the imagined velocity is 10 steps ahead relative to the prompted velocity. (**B**) 3D topographic maps of DILATE loss values for predicted and prompted velocities. The parameter *a* and *b* represent the weights and offsets of imaginary velocities mapped to predicted velocities. (**C**) Same as (B) but for MSE loss.

**Supplementary Fig. 2.**
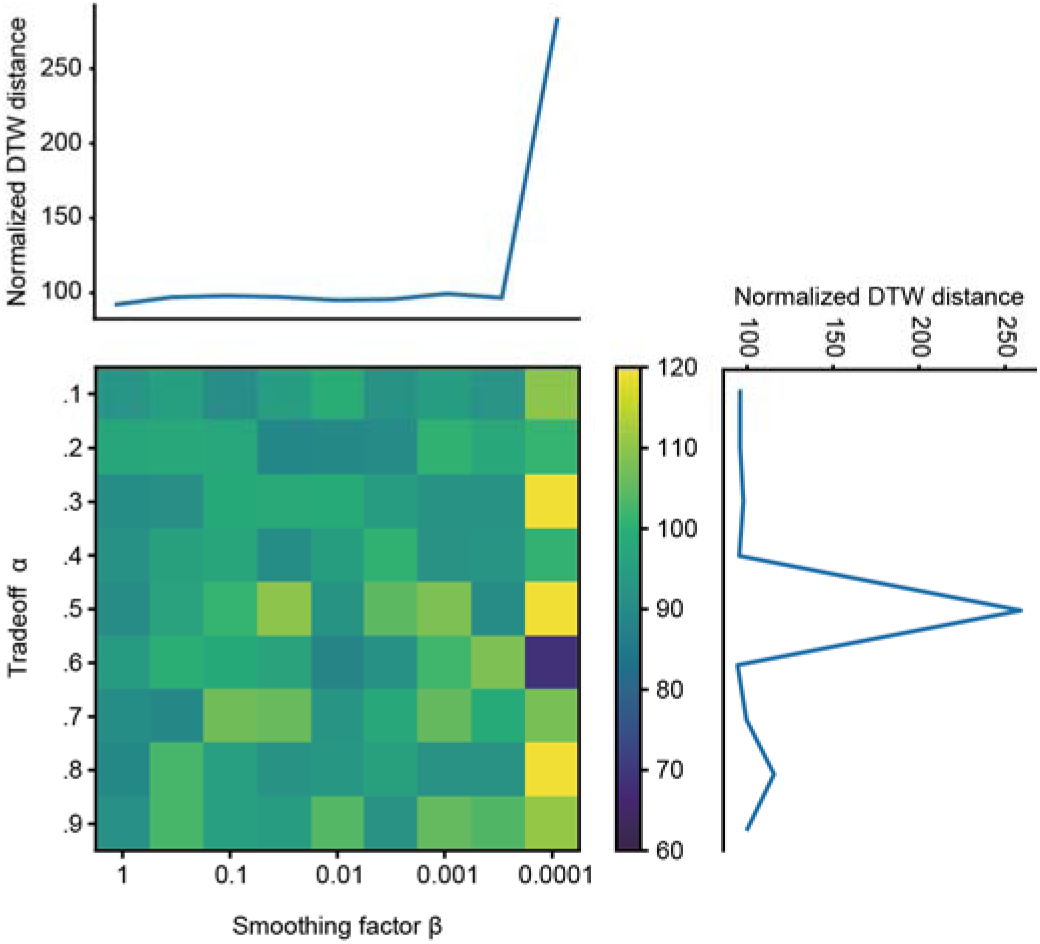
Decoding results (normalized DTW distance) for different combinations of trade-off parameter α and smoothing parameter γ. The result was obtained from one example session.

**Supplementary Figure 3.**
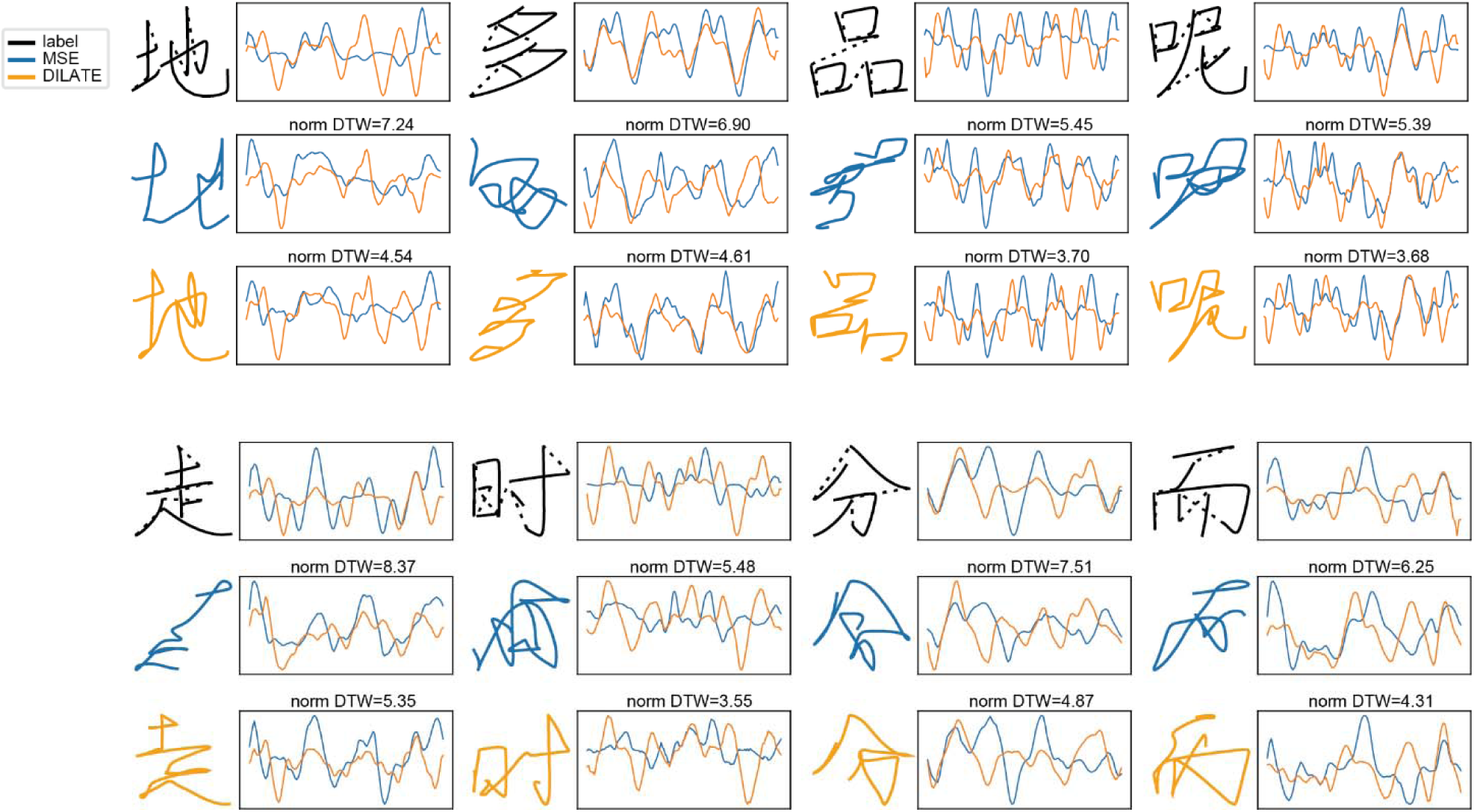
Examples of handwriting trajectory decoding with MSE and DILATE loss. The thin traces in the right side are the velocity profiles in *x-* and *y-*direction. The normalized DTW distances are labeled for each decoding.

## Notes

### Competing Interest Statement

The authors have declared no competing interest.

### Funding Statement

This work was funded by STI 2030-Major Projects (2021ZD0200404), National Natural Science Foundation of China (62336007), Pioneer R&D Program of Zhejiang (2024C03001), the Starry Night Science Fund of Zhejiang University Shanghai Institute for Advanced Study (SN-ZJU-SIAS-002), and the Fundamental Research Funds for the Central Universities (2023ZFJH01-01, 2024ZFJH01-01). The authors thank Mr. Xiang Li for software development, Prof. Schwartz for implantation surgery.

### Author Declarations

Approved by the Medical Ethics Committee of The Second Affiliated Hospital of Zhejiang University (Ethical review number 2019-158, approved on 05/22/2019), registered in the Chinese Clinical Trial Registry (chictr.org.cn; registration number: ChiCTR2100050705)

### Summary of Updates

Combine old Fig.3 and 5 as new Fig. 3; Add new Fig.2, 4, 5 and 6. Remove old Fig. 2 and 4.

